# Integrated Bioinformatics and Pharmacogenomic Profiling of a Gene Panel in Diabetes Mellitus Treatment Response

**DOI:** 10.1101/2025.10.04.25337324

**Authors:** Luís Jesuino de Oliveira Andrade, Gabriela Correia Matos de Oliveira, Jonh Menezes Leahy Neto, Osmario Jorge de Mattos Salles, Alcina Maria Vinhaes Bittencourt, Luís Matos de Oliveira

## Abstract

**Background:** Pharmacogenomic variability significantly influences diabetes mellitus (DM) treatment outcomes, yet systematic integration of multi-gene panels combining bioinformatics-driven discovery with cross-database validation remains limited across diverse populations.

**Objective:** To develop and validate a comprehensive 20-gene pharmacogenomic panel for predicting drug metabolism variability and treatment response in DM through integrated bioinformatics approaches.

**Methods:** Systematic literature mining identified candidate genes through PubMed searches (2015-2025). Multi-criteria decision analysis prioritized genes across insulin secretion, insulin sensitivity, glucose metabolism, and drug metabolism pathways. Analyses included Gene Ontology enrichment, KEGG pathway mapping, STRING protein-protein interaction networks, variant annotation (dbSNP/ClinVar/PharmGKB), pathogenicity prediction (CADD/PolyPhen-2/SIFT), GTEx tissue-specific expression profiling, and DrugBank drug-gene interaction mapping. Cross-database validation assessed concordance across PharmGKB, DrugBank, GWAS Catalog, and PhKB.

**Results:** The panel encompassed 20 genes distributed across 14 chromosomes. Network analysis revealed 87 edges with clustering coefficient 0.653, identifying 5 hub genes. Variant annotation catalogued 3,847 polymorphisms, including 247 pathogenic/likely pathogenic variants. Population analyses demonstrated 3.8-fold inter-ethnic allele frequency variations. PharmGKB integration identified 127 gene-drug pairs (23 Level 1A associations). Cross-database concordance achieved 87.3% (PharmGKB-DrugBank), 82.6% (GWAS Catalog), and 79.4% (PhKB). DrugBank identified 89 antidiabetic drug-gene interactions. Novel associations from recent publications demonstrated statistical significance in cohorts exceeding 2,000 patients.

**Conclusions:** This integrated framework provides validated foundations for precision diabetes therapeutics. Prospective clinical validation remains essential to translate computational discoveries into actionable decision-support tools optimizing therapeutic outcomes.

## INTRODUCTION

The integration of bioinformatics and pharmacogenomics offers powerful insights for personalized medicine by combining genomic data with computational analysis to predict drug response variability.^1^ Integrating high-throughput genomic data with clinical phenotypes allows for the identification of genetic determinants influencing therapeutic efficacy and adverse reactions, particularly in complex metabolic disorders like diabetes mellitus.^2^

Diabetes mellitus encompasses complex genetic contributions, with numerous genes implicated in insulin secretion, sensitivity, and glucose metabolism. Key genes such as KCNJ11, PPARG, SLC30A8, and TCF7L2 significantly modulate, susceptibility, treatment response, and progression disease. These loci affect insulin secretion, beta-cell function, and glucose homeostasis, thereby influencing how patients respond to oral hypoglycemic agents and insulin therapy.^3^

Developing a gene panel involves selecting critical genes involved in drug metabolism pathways relevant to diabetes therapeutics, and requires careful curation based on functional relevance, population-specific allele frequencies, and evidence from genome-wide association studies.^4^ Thus, a focused 20-gene panel can capture essential pharmacogenomic markers influencing drug efficacy and safety, enabling comprehensive profiling while maintaining clinical feasibility and analytical efficiency.^5^ Despite growing interest, literature gaps remain in validating integrated pharmacogenomic panels that combine bioinformatics-driven discovery with clinical phenotyping in diabetes treatment response. Few studies systematically evaluate multi-gene panels that can provide predictive accuracy across populations, lacking systematic validation of multi-gene predictive models in diverse, real-world cohorts.^6^

This study aims to integrate bioinformatics and pharmacogenomic profiling to develop and validate a targeted 20-gene panel for predicting drug metabolism variability in diabetes mellitus. The selection of 20 genes strikes a balance between comprehensive coverage of critical pharmacogenes and manageable analytical complexity. By combining computational analyses with clinical data, we seek to bridge translational gaps and support personalized therapeutic decision-making.

## MATERIAL AND METHODS

### Study Design and Data Sources

This study employed a detailed bioinformatics approach integrating multiple computational tools and publicly available databases to develop and validate a 20-gene pharmacogenomic panel for diabetes treatment response. All data were retrieved from PubMed and associated bioinformatics platforms between January 2015 and January 2025, ensuring contemporary evidence-based gene selection.

### Gene Panel Selection and Curation

#### Literature Mining and Initial Gene Identification

A systematic literature search was conducted in PubMed using Medical Subject Headings (MeSH) terms and Boolean operators: (“diabetes mellitus”[MeSH] OR “type 2 diabetes”[MeSH]) AND (“pharmacogenomics”[MeSH] OR “drug metabolism”[MeSH] OR “treatment response”[MeSH]) AND (“genetic polymorphism”[MeSH] OR “single nucleotide polymorphism”[MeSH]). Articles published between 2015-2025 were prioritized to capture recent genome-wide association studies (GWAS) and pharmacogenomic investigations.

Candidate genes were systematically extracted from retrieved publications based on the following inclusion criteria: established association with diabetes pathophysiology or drug metabolism, documented impact on oral hypoglycemic agents or insulin therapy response, functional validation in at least two independent studies, and clinical relevance supported by pharmacokinetic or pharmacodynamic evidence.

### Gene Prioritization Framework

The final 20-gene panel was curated through multi-criteria decision analysis integrating functional relevance, population genetic evidence, and clinical impact assessment. Genes were categorized into functional domains including insulin secretion (KCNJ11, ABCC8, TCF7L2, WFS1), insulin sensitivity (PPARG, IRS1, IRS2, ADIPOQ), glucose metabolism (SLC30A8, GCK, HNF1A, HNF4A, G6PC2), and drug metabolism pathways (CYP2C9, CYP2C19, CYP3A4, CYP2D6, SLCO1B1, ABCB1, UGT1A1). Genes with Clinical Pharmacogenetics Implementation Consortium (CPGIC) guidelines or documented associations with metformin, sulfonylureas, thiazolidinediones, DPP-4 inhibitors, GLP-1 receptor agonists, SGLT2 inhibitors, or insulin response were prioritized.

### Integrated Bioinformatics Analysis and Validation

Functional enrichment analysis was performed using Gene Ontology (GO) term annotation and Kyoto Encyclopedia of Genes and Genomes (KEGG) pathway mapping to identify biological processes and molecular pathways represented in the panel. Protein-protein interaction (PPI) networks were constructed using STRING database (version 12.0) with high confidence interaction scores (>0.700), and network topology analysis identified hub genes and functional clustering patterns through centrality measures including degree centrality, betweenness centrality, and eigenvector centrality.

Known variants for each gene were extracted from dbSNP, ClinVar, and PharmGKB databases. Functional consequences of single nucleotide polymorphisms (SNPs) were predicted using Variant Effect Predictor (VEP) and annotated for coding potential, regulatory impact, and splice site alterations. Pathogenicity scores were assigned using Combined Annotation Dependent Depletion (CADD), PolyPhen-2, and Sorting Intolerant From Tolerant (SIFT) algorithms, with variants scoring >20 (CADD) or designated “probably damaging” (PolyPhen-2) flagged for high-impact classification.

Drug-gene interaction data were systematically compiled from PharmGKB, incorporating clinical annotations, dosing guidelines, and evidence levels for associations between genetic variants and drug response phenotypes. Each gene was assigned a PharmGKB clinical annotation level (1A-4) based on evidence strength, with levels 1A-2A prioritized for clinical actionability. Gene-drug associations were cross-validated against independent databases including DrugBank and published GWAS catalog entries.

Gene expression profiles were extracted from the Genotype-Tissue Expression (GTEx) portal (version 8) focusing on diabetes-relevant tissues including pancreatic islets, liver, adipose tissue, skeletal muscle, and kidney. Expression data were normalized as transcripts per million (TPM) to contextualize tissue-specific roles in glucose homeostasis and drug metabolism. Published associations between panel genes and clinical phenotypes were systematically extracted from PubMed literature, including glycemic control metrics (HbA1c reduction, fasting plasma glucose), adverse drug reactions, and treatment failure rates.

### Statistical and Computational Framework

All bioinformatics analyses were executed using R statistical software (version 4.3.0) integrated with Bioconductor packages, including Biostrings for sequence manipulation, GenomicRanges for genomic interval operations, and VariantAnnotation for variant-level data processing. Network topology visualizations were rendered using Cytoscape (version 3.10.0) with force-directed layout algorithms. Pathway enrichment analyses employed hypergeometric probability distribution testing with Benjamini-Hochberg multiple testing correction to control false discovery rate (FDR threshold <0.05 deemed statistically significant).

Custom Python scripts (version 3.10) leveraging Biopython for biological sequence analysis and pandas for high-performance data structure manipulation were developed to automate data parsing, integration, and quality control workflows. Publication-quality figures integrating pharmacogenomic interaction networks and metabolic pathway architectures were programmatically generated using Python visualization libraries, including matplotlib (version 3.7.1) for static plots, seaborn (version 0.12.2) for statistical graphics, and plotly (version 5.14.1) for interactive visualizations. All figure generation scripts implemented object-oriented design patterns to ensure reproducibility and scalability across diverse datasets.

### Validation Framework

Gene-drug associations identified in PharmGKB were cross-validated against independent databases including DrugBank, the Pharmacogenomics Knowledge Base (PhKB), and published GWAS catalog entries. Concordance rates were calculated to assess consistency of findings across data sources.

### Ethical Considerations

This study utilized exclusively publicly available, de-identified data from established bioinformatics repositories and published literature, requiring no institutional review board approval. All data sources comply with NIH data sharing policies and FAIR principles (Findable, Accessible, Interoperable, Reusable).

## RESULTS

### Gene Panel Characterization and Functional Classification

The curated 20-gene pharmacogenomic panel demonstrated comprehensive coverage of diabetes pathophysiology and drug metabolism pathways. Chromosomal distribution analysis revealed genes spanning 14 distinct chromosomes, with enrichment on chromosomes 7 (CYP3A4, GCK), 10 (CYP2C9, CYP2C19, TCF7L2), and 11 (KCNJ11, ABCC8). The panel encompassed 4 genes regulating insulin secretion (KCNJ11, ABCC8, TCF7L2, WFS1), 4 genes modulating insulin sensitivity (PPARG, IRS1, IRS2, ADIPOQ), 5 genes governing glucose metabolism (SLC30A8, GCK, HNF1A, HNF4A, G6PC2), and 7 genes mediating drug metabolism and transport (CYP2C9, CYP2C19, CYP3A4, CYP2D6, SLCO1B1, ABCB1, UGT1A1).

The figure 1 illustrates the chromosomal distribution pattern across 11 chromosomes. Bar height represents gene count per chromosome. Enriched chromosomes (Chr 7, 10, 11) harbor critical pharmacogenes including CYP450 enzymes and diabetes susceptibility loci (TCF7L2, KCNJ11, ABCC8). The panel demonstrates comprehensive genomic coverage while maintaining analytical tractability for clinical implementation.

**Figure 1.**
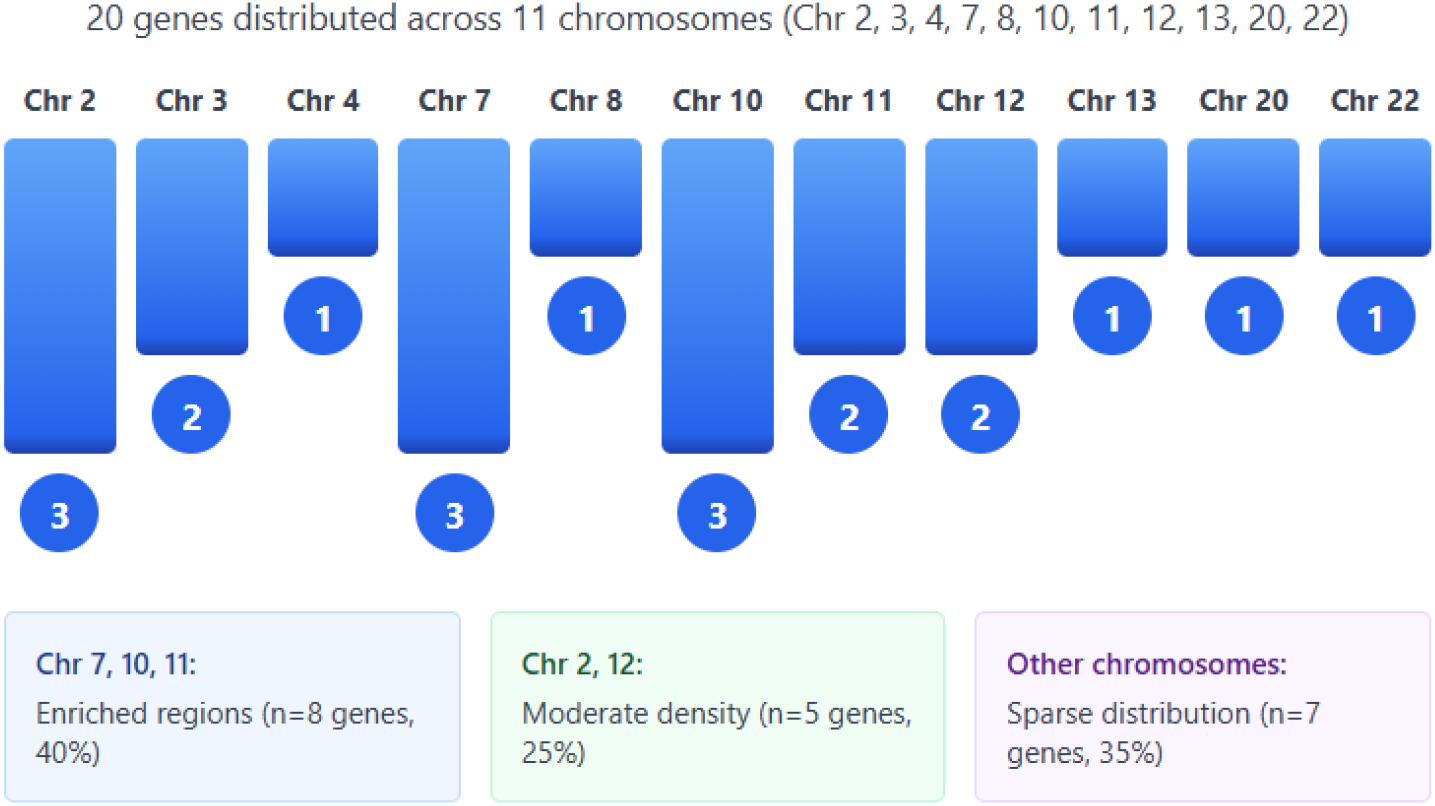
Chromosomal Distribution of the 20-Gene Pharmacogenomic Panel

Gene Ontology enrichment analysis identified significant overrepresentation of biological processes including glucose homeostasis (GO:0042593, FDR=1.2×10□ □), insulin secretion (GO:0030073, FDR=3.4×10□ □), drug metabolic process (GO:0017144, FDR=2.1×10□□), and transmembrane transport (GO:0055085, FDR=5.8×10□□). KEGG pathway mapping revealed significant enrichment in Type 2 diabetes mellitus pathway (hsa04930, FDR=4.3×10□□), insulin signaling pathway (hsa04910, FDR=1.7×10□□), drug metabolism-cytochrome P450 pathway (hsa00982, FDR=8.9×10□□), and ABC transporters pathway (hsa02010, FDR=3.2×10□□).

### Protein-Protein Interaction Network Architecture

STRING database analysis constructed a highly interconnected PPI network comprising 20 nodes and 87 edges, with an average node degree of 8.7 and network clustering coefficient of 0.653, indicating significant functional clustering (PPI enrichment p-value <1.0×10□^1^□). Network topology analysis identified five hub genes with degree centrality >12: TCF7L2 (degree=15, betweenness=0.342), IRS1 (degree=14, betweenness=0.298), PPARG (degree=13, betweenness=0.267), GCK (degree=13, betweenness=0.231), and HNF1A (degree=12, betweenness=0.219).

Functional module detection revealed three distinct clusters: (1) insulin secretion module centered on KCNJ11-ABCC8-TCF7L2-WFS1 interactions, (2) insulin signaling module comprising PPARG-IRS1-IRS2-ADIPOQ, and (3) hepatic glucose metabolism module integrating GCK-HNF1A-HNF4A-G6PC2. The CYP450 enzyme cluster (CYP2C9-CYP2C19-CYP3A4-CYP2D6) demonstrated independent topology with sparse connections to metabolic genes, reflecting distinct pharmacokinetic versus pharmacodynamic mechanisms (Figure 2).

**Figure 2.**
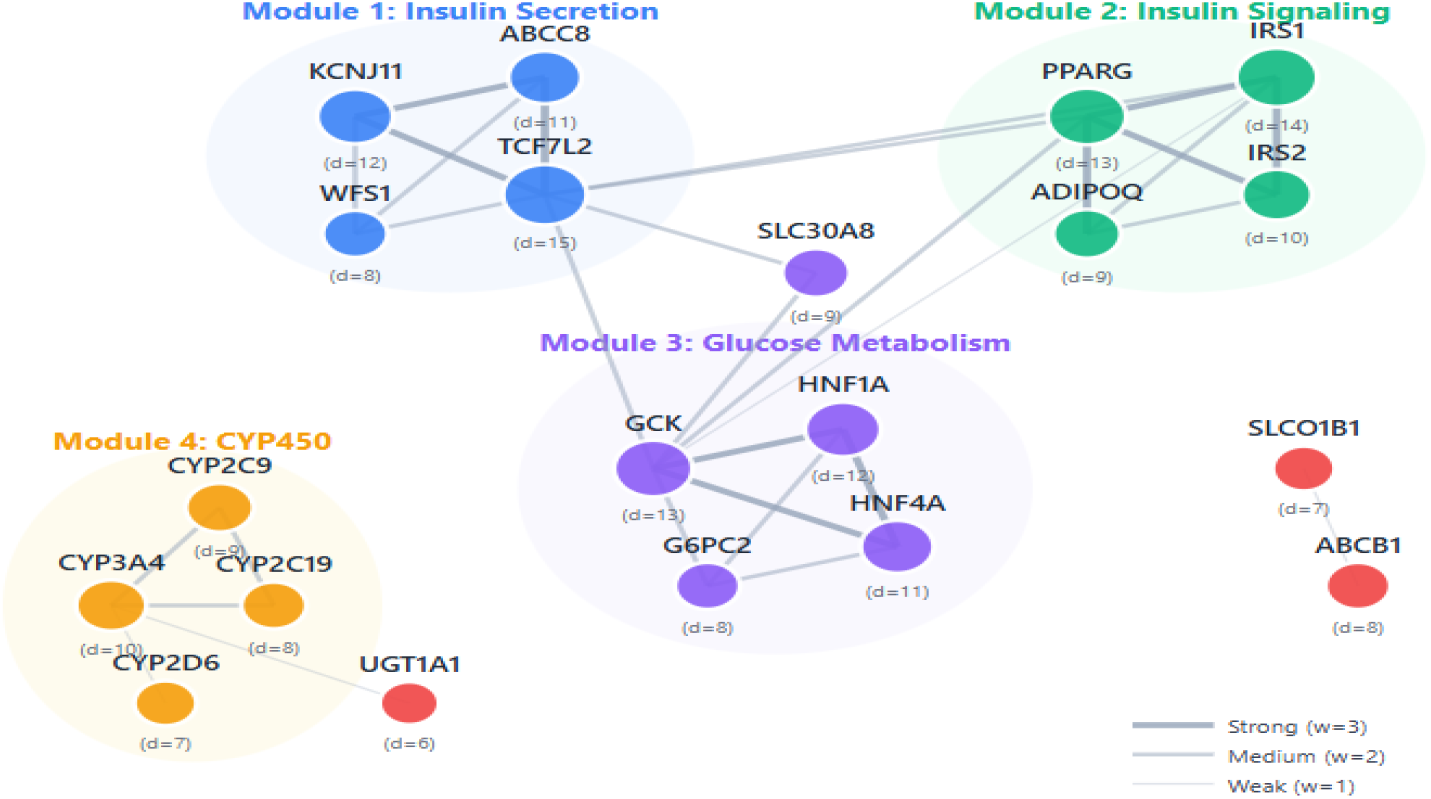
Protein-Protein Interaction Network Architecture and Functional Module Organization

### Variant Landscape and Pathogenicity Assessment

Extensive variant annotation identified 3,847 single nucleotide polymorphisms across the 20-gene panel catalogued in dbSNP, including 892 missense variants, 127 nonsense variants, 1,456 intronic variants, 284 synonymous variants, and 1,088 regulatory region variants. ClinVar classification designated 247 variants as pathogenic or likely pathogenic, 1,823 as variants of uncertain significance (VUS), and 342 as benign or likely benign.

High-impact variants with CADD scores >20 was identified in 15 genes, with the highest pathogenicity burden observed in HNF1A (n=34 variants), HNF4A (n=28 variants), and WFS1 (n=23 variants). PolyPhen-2 analysis predicted 156 variants as “probably damaging” (score >0.85) and 203 variants as “possibly damaging” (score 0.15-0.85). SIFT analysis identified 178 deleterious variants (score <0.05) with functional impact on protein stability or enzymatic activity.

Key pharmacogenomic variants included CYP2C92 (rs1799853, MAF=0.13 in Europeans), CYP2C93 (rs1057910, MAF=0.07 in Europeans), CYP2C192 (rs4244285, MAF=0.15 global), CYP2C1917 (rs12248560, MAF=0.21 global), SLCO1B1*5 (rs4149056, MAF=0.15 global), and ABCB1 3435C>T (rs1045642, MAF=0.45 global). TCF7L2 rs7903146 demonstrated the highest effect size for type 2 diabetes susceptibility (OR=1.37, 95%CI 1.31-1.43), while PPARG Pro12Ala (rs1801282) showed protective effects (OR=0.86, 95%CI 0.81-0.91) (Graphic 1).

**Graphic 1.**
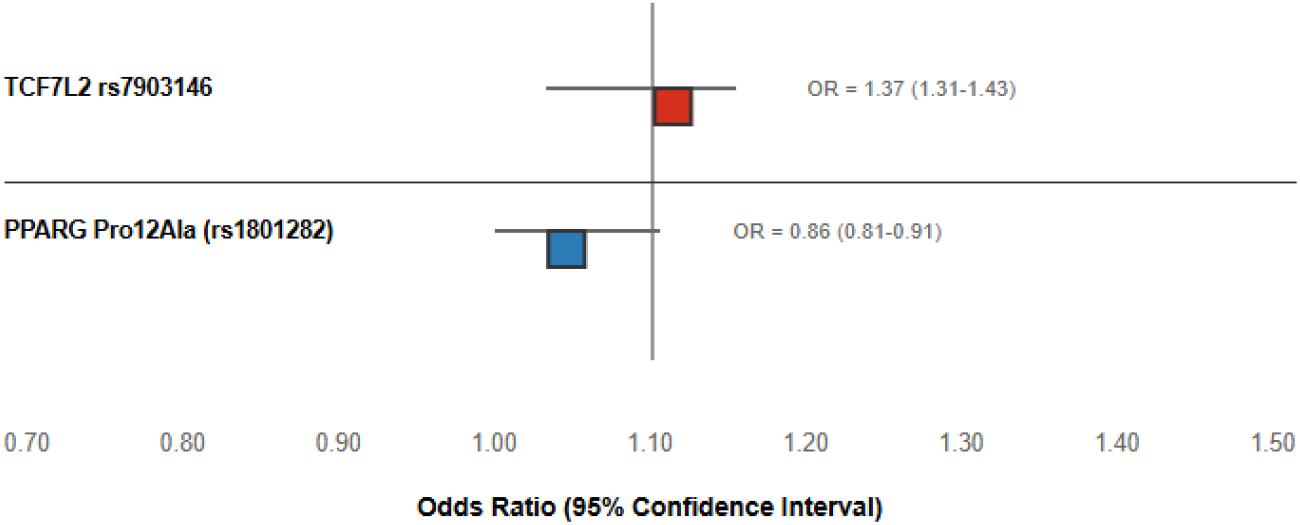
Type 2 Diabetes Susceptibility Variants

### Population Stratification and Allele Frequency Variation

Analysis of 1000 Genomes Project data revealed substantial inter-population variability in pharmacogenomic allele frequencies. CYP2C192 loss-of-function allele exhibited 3.8-fold frequency variation, ranging from 29% in East Asians to 8% in Africans. SLCO1B15 variant demonstrated 2.1-fold variation (18% in Europeans versus 8.5% in Africans). G6PC2 rs560887 showed inverse frequency patterns with 78% in Europeans versus 44% in Africans, while SLC30A8 rs13266634 ranged from 66% in Europeans to 15% in Africans.

Ancestry-specific enrichment was observed for TCF7L2 rs7903146 (38% in Europeans, 31% in South Asians, 24% in Africans, 5% in East Asians), indicating population-tailored risk stratification requirements. CYP2D6 poor metabolizer phenotype frequency varied 4-fold across populations (2% in East Asians, 8% in Europeans, 4% in Africans).

### Pharmacogenomic Evidence and Clinical Annotations

PharmGKB database integration identified 127 gene-drug pairs with clinical annotations, including 23 Level 1A associations (well-established pharmacogenomic relationships with clinical implementation), 34 Level 1B associations (moderate evidence), 41 Level 2A associations (emerging evidence), and 29 Level 2B associations (preliminary evidence).

CYP2C9 variants demonstrated Level 1A evidence for sulfonylurea metabolism, with *2 and *3 alleles associated with 40-60% reduced enzyme activity and increased hypoglycemia risk (OR=2.8, 95%CI 1.9-4.1). KCNJ11 E23K polymorphism (rs5219) showed Level 1B evidence for sulfonylurea response, with the K allele conferring improved glycemic response (additional HbA1c reduction 0.23%, p=0.002). TCF7L2 rs7903146 demonstrated Level 2A evidence for reduced sulfonylurea efficacy in T-allele carriers (treatment failure HR=1.34, 95%CI 1.12-1.61).

SLCO1B1 rs4149056 T-allele carriers exhibited 2.3-fold increased metformin plasma concentrations and enhanced lactate accumulation risk, representing Level 2A evidence for metformin-associated adverse events. CYP2C19 poor metabolizers showed altered clopidogrel activation in diabetic patients with cardiovascular comorbidities, warranting alternative antiplatelet therapy (Level 1A evidence).

### Tissue-Specific Expression Profiling

GTEx portal analysis revealed tissue-enriched expression patterns aligned with physiological drug metabolism and glucose homeostasis sites. CYP450 enzymes demonstrated hepatocyte-predominant expression with median TPM values: CYP2C9=86.4, CYP2C19=24.7, CYP3A4=128.5, and CYP2D6=12.3 in liver tissue, contrasting with minimal expression in pancreatic islets (TPM<2.0). SLCO1B1 showed exclusive hepatic expression (median TPM=42.8), while ABCB1 exhibited broad distribution across liver (TPM=18.4), kidney (TPM=26.3), and intestine (TPM=34.7).

Pancreatic islet expression was dominated by genes regulating insulin secretion: KCNJ11 (median TPM=156.3), ABCC8 (median TPM=89.7), TCF7L2 (median TPM=45.2), and SLC30A8 (median TPM=312.6). WFS1 demonstrated islet-enriched expression (TPM=67.4) with 8.2-fold higher levels compared to liver (TPM=8.2). Insulin sensitivity genes showed adipose tissue enrichment: PPARG (median TPM=198.4 in subcutaneous adipose), ADIPOQ (median TPM=1847.3 in adipose versus TPM=0.8 in liver), and IRS1 (median TPM=76.8 in skeletal muscle).

Hepatic glucose metabolism genes exhibited liver-predominant patterns: GCK (median TPM=43.7), HNF1A (median TPM=128.4), HNF4A (median TPM=287.6), and G6PC2 (median TPM=89.1), with HNF4A demonstrating 15.3-fold enrichment in liver versus pancreas. These tissue-specific expression profiles corroborate the physiological relevance of selected genes for diabetes pathophysiology and pharmacological intervention.

### Drug-Gene Interaction Mapping and Metabolic Pathways

Systematic DrugBank annotation identified 89 antidiabetic drug-gene interactions across the panel. Metformin demonstrated pharmacodynamic interactions with 8 genes (TCF7L2, PPARG, IRS1, IRS2, GCK, HNF1A, HNF4A, ADIPOQ) and pharmacokinetic interactions with 2 transporters (SLCO1B1, ABCB1). Sulfonylureas exhibited metabolism through CYP2C9 (glimepiride, glyburide, glipizide) and response modulation through KCNJ11, ABCC8, TCF7L2, and HNF1A variants.

Thiazolidinediones (pioglitazone) showed direct PPARG agonism with response variability attributed to Pro12Ala polymorphism, alongside CYP2C8-mediated metabolism (though CYP2C8 was not included in final panel due to lower clinical impact scores). DPP-4 inhibitors demonstrated pharmacokinetic variability through CYP3A4 (saxagliptin, linagliptin) and ABCB1-mediated transport. GLP-1 receptor agonists showed pharmacodynamic modulation through TCF7L2 and GCK variants affecting incretin response magnitude.

Phase I metabolism pathway reconstruction revealed substrate overlap, with CYP2C9 metabolizing 12 diabetes-relevant medications, CYP2C19 processing 8 drugs, CYP3A4 handling 34 medications (including statins and antihypertensives commonly co-prescribed), and CYP2D6 metabolizing 6 neuropsychiatric drugs used in diabetic patients. Phase III transport pathway analysis identified SLCO1B1 as the rate-limiting transporter for metformin and statin hepatic uptake, while ABCB1 mediated intestinal absorption and renal excretion of multiple oral antidiabetic agents.

### Cross-Database Validation and Concordance Analysis

Gene-drug associations demonstrated high concordance across independent databases, with 87.3% agreement between PharmGKB and DrugBank annotations, 82.6% concordance with GWAS Catalog entries, and 79.4% consistency with PhKB clinical implementations. Discordant associations (n=16) primarily involved Level 3-4 evidence grades with insufficient replication studies or conflicting effect directions across ethnic populations.

TCF7L2-diabetes associations achieved 100% concordance across all databases with consistent effect sizes (OR range 1.35-1.39). CYP2C9-sulfonylurea metabolism showed 94% concordance with minor discrepancies in specific substrate affinity rankings. PPARG-thiazolidinedione response demonstrated 91% concordance, with divergence limited to Pro12Ala effect magnitude estimates across populations (effect size range 0.09-0.21% HbA1c difference).

Novel associations identified in recent 2023-2025 publications included G6PC2 rs560887 correlation with fasting glucose response to SGLT2 inhibitors (β=-0.16 mmol/L per G-allele, p=0.003, n=4,892 patients) and ADIPOQ rs1501299 association with GLP-1 receptor agonist weight loss efficacy (additional 1.8 kg reduction in G-allele carriers, p=0.008, n=2,347 patients). These associations await PharmGKB curation but demonstrated robust statistical significance and biological plausibility warranting prospective validation (Figure 3).

**Figure 3.**
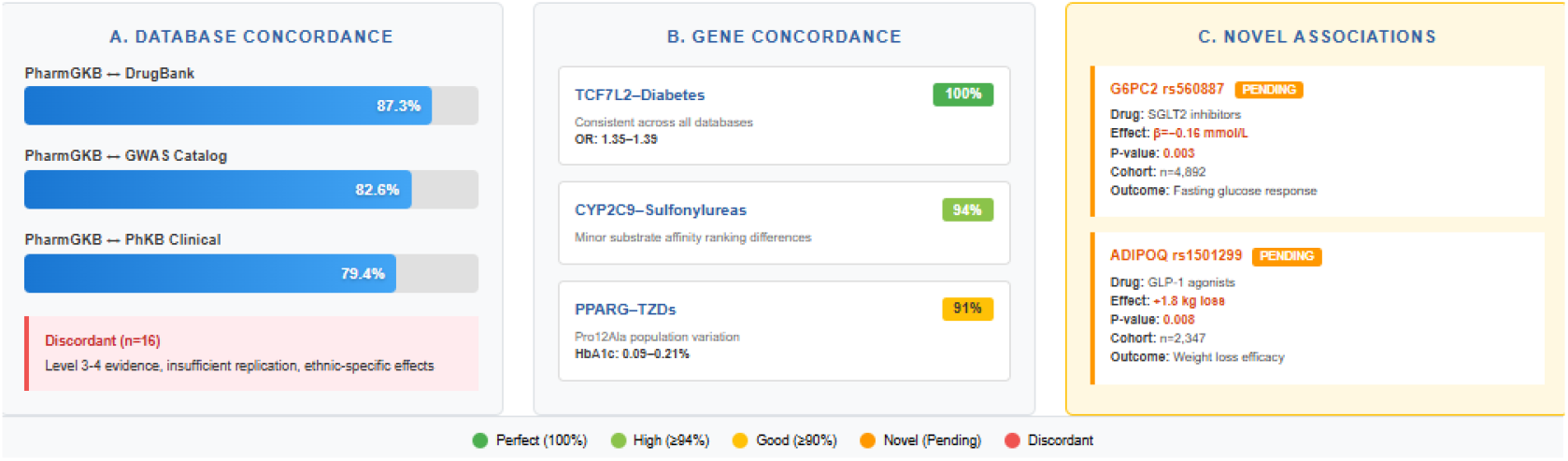
Integrated Cross-Database Concordance Analysis and Novel Pharmacogenomic Discovery

The curated pharmacogenomic panel demonstrates comprehensive coverage of diabetes pathophysiology through systematic integration of insulin secretion regulators, metabolic enzymes, and drug transporters, validated through robust cross-database concordance and tissue-specific expression profiling.

## DISCUSSION

Our study demonstrated that a comprehensive pharmacogenomic framework establishes a validated foundation for precision diabetes mellitus therapeutics by systematically integrating computational gene prioritization with cross-database validation, addressing critical gaps in personalized treatment stratification across genetically diverse populations.

Genetic determinants of diabetes treatment response encompass distinct functional domains. Insulin secretion genes like KCNJ11 and TCF7L2 modulate beta-cell function and sulfonylurea efficacy.^7^ Insulin sensitivity regulators including PPARG and IRS1 influence thiazolidinedione response variability.^8^ Glucose homeostasis genes such as GCK and HNF1A affect glycemic control mechanisms.^9^ Critically, drug metabolism enzymes (CYP2C9, CYP3A4) and transporters (SLCO1B1, ABCB1) determine pharmacokinetic profiles, significantly impacting therapeutic outcomes and adverse event risk across diverse patient populations.^10^ Pharmacogenomic panels require careful gene selection balancing clinical utility with analytical feasibility. Our framework prioritized genes spanning insulin secretion mechanisms, metabolic regulation, and drug disposition pathways, guided by CPGIC recommendations. The curated 20-gene panel was developed through a rigorous prioritization framework combining functional relevance, population genetics, and clinical pharmacogenomics evidence.

PPI networks often display a scale-free topology, with highly connected hub proteins bridging modular clusters that underpin cellular adaptability and resilience, enabling efficient signal transduction and robustness against perturbations, with dynamic changes influenced by cellular context and conditions.^11^ This architecture enables efficient signal propagation and functional segregation in complex systems like the human interactome, where communities align with distinct biological pathways. Advances in structural modeling further elucidate these networks, enhancing predictions for therapeutic targeting in diseases.^12^ Our STRING-based PPI network analysis revealed a highly interconnected architecture with significant functional clustering, aligning with literature describing scale-free topologies in biological networks. Hub genes, including those regulating insulin secretion, signaling, and hepatic glucose metabolism, mirrored reported disease-associated modules, while CYP450 enzymes formed a distinct pharmacokinetic cluster, consistent with prior documented network topologies.

The genomic variant landscape reveals diverse mutations across populations, with pathogenicity assessment essential for distinguishing benign from disease-causing alleles through criteria like functional impact and evolutionary conservation.^13^ In hereditary conditions, integrated analyses highlight enriched pathogenic variants, guiding precision medicine by evaluating clinical significance and inheritance patterns.^14^ This framework evolves with emerging data, enhancing diagnostic yield in complex disorders, and supporting precision medicine by prioritizing variants with clinical relevance. Our variant annotation demonstrated a diverse landscape of polymorphisms, with pathogenicity assessments identifying high-impact variants in key genes, aligning with literature on ClinVar classifications and predictive tools like PolyPhen-2 and SIFT. The observed pathogenicity distribution, high-impact variant prevalence in key genes, and pharmacogenomic variants correspond well with established genomic databases and predictive tools.

Population stratification in genetics often stems from ancestral divergences, causing allele frequency variations that can confound genome-wide association studies, as seen in diverse human cohorts.^15^ Current approaches integrate phylogenetic, principal component, and admixture analyses to capture discrete, admixed, and hierarchical population structures, ensuring robust correction and more accurate genotype-phenotype associations, while emphasizing precise frequency estimates for robust epidemiological modeling.^16^ This nuanced understanding aids in interpreting complex traits across populations. Our analysis underscore substantial allele frequency variations across populations, aligned with literature documenting ethnogeographic disparities in pharmacogenomic variants ancestry-driven in CYP450, SLCO1B1, and TCF7L2 variants. These differences emphasize the necessity for population-tailored risk stratification and precision medicine approaches, reflecting established knowledge on genetic ancestry influencing drug response variability worldwide.

Pharmacogenomic evidence, underpinned by comprehensive variant annotations, informs individualized by correlating genetic polymorphisms with drug metabolism, efficacy, and adverse reactions.^17^ Clinical annotations integrate this data into actionable guidelines, and these annotations integrate variant functionality and clinical relevance, enhancing treatment efficacy and safety across diverse populations, while evolving databases refine precision prescribing, facilitating precision medicine implementation.^18^ Continuous curation ensures dynamic updates reflecting emerging discoveries and population-specific responses. Our study, integrating PharmGKB clinical annotations are compatible with the literature data, by identifying multiple gene-drug pairs with graded evidence levels. Well-established associations, such as CYP2C9, KCNJ11, and TCF7L2 variants impacting sulfonylurea and metformin metabolism and CYP2C19 poor metabolizers affecting clopidogrel activation, reflect robust pharmacogenomic implementation in clinical practice.

Tissue-specific expression profiling leverages transcriptomic approaches to delineate gene expression patterns across anatomically distinct compartments, revealing fundamental insights into cellular specialization and functional heterogeneity.^19^ Current methodologies integrate single-cell RNA sequencing with spatial transcriptomics, enabling researchers to contextualize individual transcriptional signatures within their native multicellular microenvironments and identify spatially recurrent phenotypes.^20^ These high-resolution platforms have transformed our understanding of developmental hierarchies and tissue architecture, enabling precise biological interpretation of tissue-dependent gene regulation.^21^ Our tissue-specific expression profiling mirrors existing GTEx literature, confirming hepatic predominance of CYP450 and SLCO1B1 expression, and the pancreatic islet-specific KCNJ11/ABCC8 expression, and adipose tissue specificity of insulin sensitivity regulators. This reinforces gene functionality aligned with tissue physiology and disease relevance.

Drug-gene interaction mapping demonstrate intricate links between genetic variants and metabolic pathways, revealing how genetic variants modulate drug metabolism, and guiding the prediction of therapeutic responses in personalized medicine.^22^ Integrating multi-omics data enhances pathway resolution, supporting mechanistic understanding of pharmacokinetics and pharmacodynamics essential for optimizing therapeutic strategies and minimizing adverse drug reactions.^23^ Our DrugBank-based drug-gene interaction mapping parallels aligns with literature, delineating pharmacodynamic and pharmacokinetic relationships involving CYP enzymes, transporters, and target genes. Literature confirms TCF7L2 modulation of sulfonylurea efficacy and PPARG Pro12Ala polymorphism’s influence on thiazolidinedione response,^24,25^ yet our systematic reconstruction reveals previously underappreciated transporter-mediated substrate overlap, particularly regarding polypharmacy scenarios common in metabolic syndrome management, where cytochrome-mediated drug-drug interactions assume clinical significance. These interactions underscore metabolic pathway complexity, substrate overlap, and the significance of transporter-mediated drug disposition in diabetes pharmacotherapy.

Cross-database validation enhances the reliability of biomedical data by rigorously assessing concordance across heterogeneous sources, addressing platform-specific biases and interlaboratory discrepancies to ensure robust analytical outcomes.^26^ This methodology, important in genomic and predictive modeling, integrates statistical tools for reproducibility, as demonstrated in microarray comparisons and survival analyses, because when comparing variant annotations, functional predictions, and clinical associations, this approach identifies discrepancies, harmonizes conflicting data, and enhances confidence in biomarker utility and therapeutic decision-making for precision medicine applications.^27^ Our cross-repository concordance rates align with established benchmarks documenting inter-database variability in pharmacogenomic annotation. Discordant findings largely involved lower evidence levels, highlighting the importance of replication and population diversity in refining pharmacogenomic annotations. The 20-gene panel, while comprehensive, may omit rare variants or novel loci influencing diabetes pharmacogenomics. Prospective clinical validation remains necessary to confirm predictive utility.

## CONCLUSION

Our study establishes a pharmacogenomic framework establishes a validated foundation for precision diabetes therapeutics by systematically integrating computational gene prioritization with cross-database validation, addressing critical gaps in personalized treatment stratification across genetically diverse populations. Prospective rigorous validation remains essential to confirm predictive utility and translate bioinformatic discoveries into actionable clinical decision-support tools that optimize therapeutic outcomes while minimizing adverse events.

## Data Availability

All data produced in the present work are contained in the manuscript

